# Health-Related Quality of Life in the Spironolactone to Reduce ICD Therapy (SPIRIT) Trial

**DOI:** 10.1101/2020.08.27.20016378

**Authors:** Ana Carolina Sauer Liberato, Merritt H. Raitt, Ignatius GE Zarraga, Karen S. MacMurdy, Cynthia M. Dougherty

**Affiliations:** Institute of Nursing, University of Freiburg, Germany; VA Portland Health Care System, Division of Cardiology, Portland, OR; Knight Cardiovascular Institute, Oregon Health and Sciences University, Portland OR; University of Washington School of Nursing, Department of Biobehavioral Nursing and Health Informatics, Seattle, WA; VA Puget Sound Health Care System, Seattle, WA

**Keywords:** Health-related quality of life, ICD, spironolactone, defibrillator, implantable

## Abstract

**Purpose:** The Spironolactone to Reduce ICD Therapy (SPIRIT) trial was designed to determine whether once daily spironolactone would: 1) reduce the incidence of ventricular tachycardia (VT) and ventricular fibrillation (VF), and 2) improve health related quality of life (HRQOL) and symptoms. The purpose of this paper is to describe the long term HRQOL outcomes in the SPIRIT trial and compare QOL in those who did or did not receive implantable cardioverter defibrillator (ICD) shocks during follow-up.

**Methods:** Ninety participants age 66±10 years, 96% men, 75% with NYHA class II, with an ICD at moderately high risk for recurrent VT/VF were randomized to spironolactone 25mg (N=44) or placebo (N=46). HRQOL was measured every 6 months for 24 months using 3 instruments: Patient Concerns Assessment (PCA), Short Form Health Survey-Veterans Version (SF-36V), and the Kansas City Cardiomyopathy Questionnaire (KCCQ). Linear mixed modeling was used to compare changes in HRQOL across 24-months. ANCOVA was used to compare HRQOL between those getting an ICD shock or not.

**Results:** Over 24-months, there were no differences in HRQOL between the spironolactone vs. placebo groups. Those who experienced at least 1 ICD shock vs. those with no ICD shocks, reported significantly lower HRQOL and more symptoms at 6-months and 24 months.

**Conclusions:** Spironolactone had no significant impact on HRQOL. Patients receiving one or more ICD shocks reported significant reductions in HRQOL and higher symptoms.

## INTRODUCTION

The use of the implantable cardioverter defibrillator (ICD) for life-threatening ventricular arrhythmias is a standard therapy, in large part because clinical trials have consistently demonstrated its superiority over medical therapy in preventing sudden cardiac death in selected patients with heart failure or prior cardiac arrest.^1,2^ In the 1990s, the Randomized Aldactone Evaluation Study (RALES) trial demonstrated that in patients with heart failure with reduced ejection fraction (HF*r*EF), aldosterone blockade was associated with a reduced risk of sudden cardiac death by 30% in those with moderate to severe HF symptoms after myocardial infarction^3,4^ and improved symptoms. In patients with heart failure with preserved ejection fraction (HF*p*EF), adding spironolactone to existing therapy did not significantly reduce death from cardiovascular causes, abort cardiac arrest, nor reduce hospitalization for heart failure.^5^ However, the use of spironolactone was associated with an improvement in HF-specific health-related quality of life (HRQOL).^6^ In our primary study, **SPI**ronolactone to **R**educe **I**CD **T**herapy (SPIRIT) Trial, we hypothesized that spironolactone at 25 mg/day would delay the first occurrence of ventricular tachycardia (VT)/ventricular fibrillation (VF) and reduce the risk of VT/VF in patients with ICDs^7^ over 24 months. We also hypothesized that spironolactone would improve HRQOL and reduce HF symptoms. The primary outcomes of the study have been reported and this hypothesis was not supported.^7^

It has been demonstrated that patients who receive ICD shocks experience reduced QOL,^8^ where an ICD shock is associated with reduced physical functioning and mental health. Others have reported that patients who experience an ICD shock do not adapt well to living with the ICD and are more anxious than those who have not received shocks.^9^ The gap in the literature that this paper addresses is the effect of an aldosterone blocker on overall HRQOL and symptoms in patients with an ICD over a longer period of time than has been previously reported, and the effect of ICD shocks on HRQOL and symptoms. Therefore, the purpose of this paper is to report the impact of long-term spironolactone administration on HRQOL and symptoms and to describe the effect of ICD shocks on HRQOL over 24-months.

## METHODS

### Design and Population

In the SPIRIT study, persons with an ICD were enrolled into a double-blind prospective stratified randomized control trial if they had received an ICD therapy, either a shock or anti-tachycardia pacing (ATP) for VT/VF in the previous 2-years or received an ICD for secondary prevention with sustained VT/VF in the previous 6 months. Additional inclusion criteria included enrollment in one of the follow-up clinics, a normally functioning ICD, and ability to speak and understand English. The exclusion criteria were: indication for spironolactone based on the Randomized Aldactone Evaluation Study (RALES)^3^ (EF of <35% and NYHA class III or IV), unstable angina, primary hepatic failure, known intolerance to spironolactone, serum creatinine concentration of >2.5 mg/dL, serum potassium concentration of >5.0 mmol/L, or a life expectancy of <2 years. Additional details about the study and the primary outcomes are published.^7^

Patients were enrolled from the Portland Veterans Affairs Medical Center, Veterans Affairs Puget Sound Health Care System, Oregon Health and Science University, and the Little Rock Veterans Affairs Medical Center. The institutional review board or ethics committee at each site approved the protocol, and all patients provided written informed consent before enrollment.

### Protocol

Patients were randomized to spironolactone at 25 mg/day or placebo based on the number of months since the last ICD shock and NYHA functional class. The Research Pharmacy service of the VA Portland Health Care System performed randomization using a computer-generated randomization scheme. The placebo and spironolactone capsules were identical in appearance. The investigators and clinicians who were involved with ICD interrogation, interpretation of arrhythmias, ICD programming, and outpatient and in-hospital management were blinded to the patients’ treatment assignment. Patients were evaluated in an outpatient setting at the time of randomization and were subsequently followed every 3 months for 24 months until the termination of the study. ICD programming (detection intervals for VT/VF and therapies) and the use of antiarrhythmic drugs were left to the discretion of the electrophysiologists who implanted the device or followed the patients in the outpatient clinic. Pill counts and pharmacy refills were used to verify adherence to spironolactone and placebo medications every three months.

### Data Collection and Outcomes

Three HRQOL questionnaires were used to measure general and disease-specific HRQOL at baseline randomization and then every 6 months for 24 months. Data were collected in the clinic before each study associated ICD clinic visit.

1. <u>Patient Concerns Assessment (PCA)</u> is a symptom checklist that measures physical symptoms and fears that are common after ICD implantation. The PCA is a disease-specific instrument for ICD QOL, symptoms, and distress, with a reliability of (α = 0.88).^10^ Higher scores reflect more concerns, fears, and symptoms.
2. <u>Short Form Health Survey adapted for veterans (SF-36V)</u> is a 36-item questionnaire that measures general physical and mental health.^11^ The SF-36V is a reliable and valid questionnaire, containing eight constructs of health status: physical functioning (PF), role limitations due to physical problems (RP), bodily pain (BP), general health perceptions (GH), energy/vitality (VT), social functioning (SF), role limitations due to emotional problems (RE), and mental health (MH). These eight dimensions can be summarized numerically into two scores, the physical component summary (PCS) and the mental component summary (MCS).^11^ Higher scores reflect better HRQOL.
3. <u>Kansas City Cardiomyopathy Questionnaire (KCCQ)</u> is a 23-item reliable and valid questionnaire,^12^ which evaluates HRQOL in heart failure. It quantifies, in a disease-specific fashion, physical limitations, symptoms, quality of life, social interference and self-efficacy.^12^ KCCQ provides the calculation of two main scores, the overall score and the clinical summary score, which includes functional status, social limitation and quality of life domains scores. For this study the Cronbach’s alpha reliability was α = 0.93. Higher scores reflect higher HRQOL.

At each clinic visit an ICD interrogation was performed, and any ICD therapy (shock or ATP) was noted. The stored intracardiac electrograms and other details of each ICD therapy were downloaded, saved, and reviewed independently by two electrophysiologists. In addition, at each visit, information about potential drug side effects and any hospitalization after randomization was obtained.

### Statistical Analysis

An intention-to-treat analyses between spironolactone and placebo groups were conducted followed by an analysis based on receipt of an ICD shock or no ICD shock in each 6-month block of time. The sample size of the study was calculated based on the primary outcome. Descriptive statistics were used to examine characteristics of the study participants (Table 1). Comparisons of baseline demographic and clinical variables between the spironolactone group and placebo groups were done using Chi-Square (χ2) tests for dichotomous variables and t-tests for continuous variables.

**Table 1.** Baseline Demographic and Clinical Characteristics

| Characteristic | Placebo<br>(n=46) | SPL<sup>†</sup><br>(n=44) | *t* or $\chi^2$ | *p* |
| --- | --- | --- | --- | --- |
| Age (years) | 66.5±9.3 | 66.6±10.9 | 0.53 | 0.96 |
| Male, n (%) | 43 (93) | 44(100) | 2.97 | 0.24 |
| Caucasian race, n (%) | 43(94) | 44(100) | 2.97 | 0.24 |
| ICD indication, n (%) Secondary | 41(89) | 36(81) | 0.97 | 0.32 |
| Number of comorbidities | 3.8± 1.4 | 4.2 ± 1.36 | 1.39 | 0.17 |
| Serum potassium, mmol/L | 4.2±0.3 | 4.3±0.4 | 0.42 | 0.94 |
| Serum creatinine, mg/dl | 1.2±0.3 | 1.2±0.3 | 0.20 | 0.99 |
| EF* (%) | 34.5 ±12.2 | 33.4 ± 13.4 | 0.40 | 0.69 |
| NYHA class, n (%) |  |  | 0.56 | 0.54 |
| I | 10(22) | 7(16) |  |  |
| II | 34(74) | 34(77) |  |  |
| III | 2(4) | 3(7) |  |  |
| IV | 0(0) | 0(0) |  |  |
| Coronary Artery Disease, n (%) | 39(85) | 41(93) | 1.61 | 0.32 |
| Myocardial Infarction, n (%) | 27(59) | 30(68) | 0.87 | 0.35 |
| Percutaneous Coronary intervention | 9(20) | 13(30) | 1.21 | 0.27 |
| Coronary Artery Bypass Graft<br>Surgery | 18(39) | 18(41) | 0.03 | 0.86 |
| Congestive Heart Failure | 32(70) | 38(83) | 3.67 | **0.05** |
| Dilated Cardiomyopathy | 6(13) | 3(7) | 0.97 | 0.49 |
| Hypertension | 28(61) | 29(66) | 0.25 | 0.62 |
| Diabetes Mellitus | 16(35) | 13(29) | 0.28 | 0.59 |
| Shocks (total number) | 6.78(14.87) | 7.93(12.03) | 0.40 | 0.69 |

\*Ejection Fraction; †Spironolactone

Outcome analyses were carried out by fitting linear mixed models by using the Statistical Package for the Social Sciences (SPSS, v 21.0). Separately for each outcome, we used generalized estimating equations (GEE) that started with a model that included fixed effects associated with group assignment, time since randomization (in months), and an interaction between those two terms. We fit each model with a random intercept; a random intercept and slope; and a random intercept, slope, and quadratic trajectory, incorporating fixed effects for quadratic patterns over time. The random slope terms allowed different individuals to have different time trends, and the random quadratic terms allowed different individuals to have different curvature in the patterns of outcomes over time. Baseline scores were included in the models to control for variations between the groups on these measures. Analysis of variance (ANOVA) controlling for the baseline value of the variable in shock (experiencing at least 1 shock before the measurement time) versus no shock groups was performed. The 95% confidence intervals and 2-sided p-values are reported. An alpha ≤0.05 was considered statistically significant. Because shock and no shock groups were created post-randomization, these findings are presented as associations and are to be interpreted more conservatively.

## RESULTS

### Study Participants

Ninety (90) patients were randomized to the spironolactone (44) or placebo group (46) and followed for 24 months. The majority of study participants were white males, with a mean age of 66.5±9.3 years for the placebo group and 66.6±10.9 years for spironolactone group (Table 1). The main reason for receipt of the ICD was secondary prevention. The placebo group received on average (mean±SD) 6.8±14.9 shocks and the spironolactone group 7.9±12.0 shocks over 24 months. None of the demographic or clinical characteristics of the patients were significantly different between placebo and spironolactone groups. Most participants were taking an ACE inhibitor, beta blocker, loop diuretic, aspirin and statin medications (Table 2). There were 20% in the placebo group and 25% in the spironolactone group taking Amiodarone (Table 2).

**Table 2.**
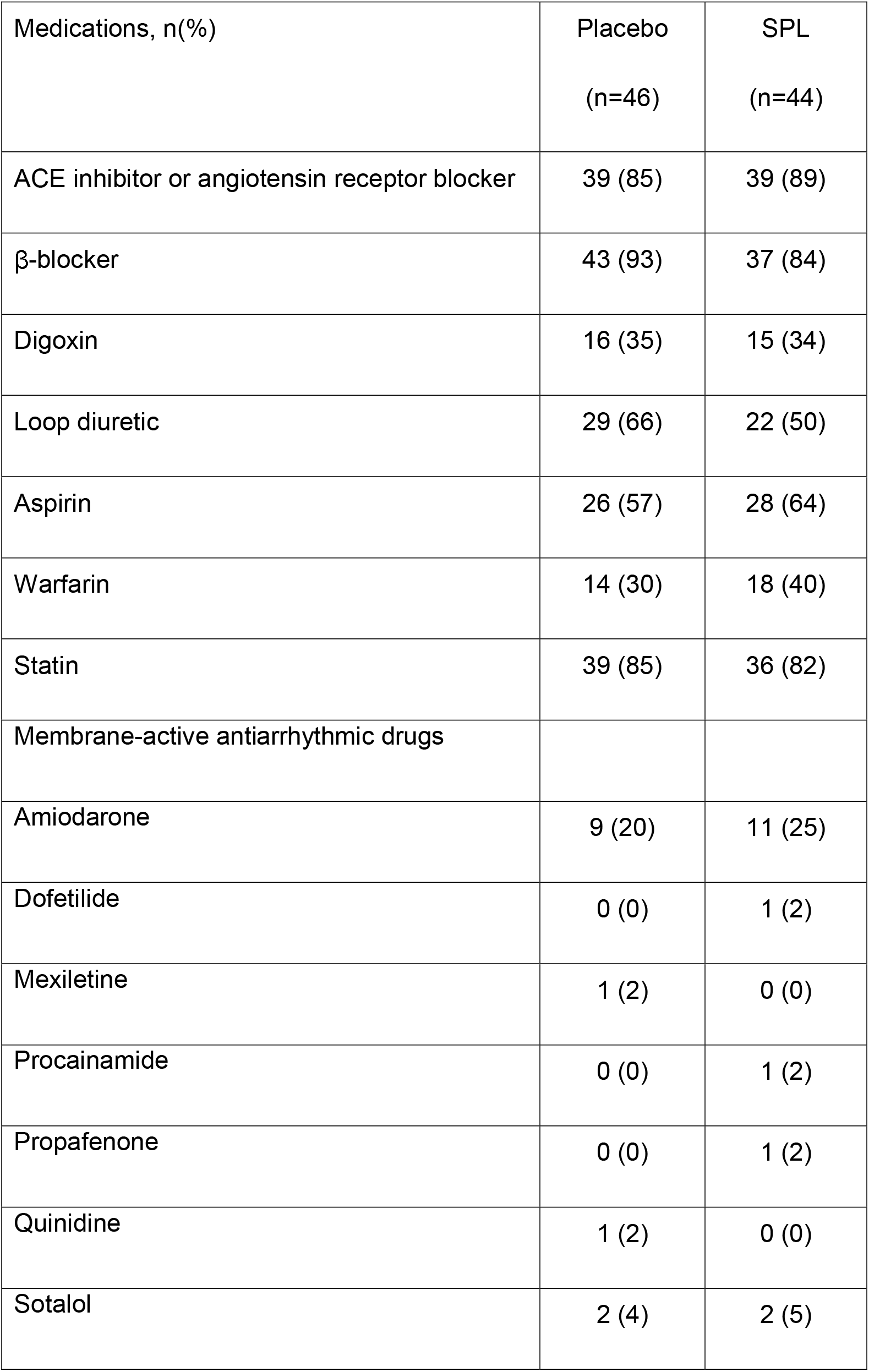
Baseline medication use

More detailed information about the SPIRIT trial design, recruitment, randomization and other clinical characteristics of the patients has been published.^7^ There were initially 44 in the spironolactone group: 30 completed the 24-months of follow-up, with 8 drop-outs and 6 deaths (Figure 1). In the placebo group there were initially 46: 30 completed the 24-months of follow-up, with 6 dropouts and 10 deaths. No patient was lost to follow-up, and no patient crossed over to the other treatment arm. During the study, 34 patients (15 in the placebo group and 19 in the spironolactone group; *p*=0.39) permanently discontinued the study medication. The most frequent reasons for discontinuation of the study medication was hyperkalemia, gynecomastia, renal dysfunction, dizziness, or diarrhea. Patient taking spironolactone reported more gynecomastia than those taking placebo (9% vs, 0%, *p*=0.05). The median time from randomization to the last dose of spironolactone was 16 months for patients in the placebo group and 7 months for those in the spironolactone group. There were no statistically significant differences in reported adverse events between the two groups, but there was a trend noted in more hospitalizations for ICD shocks, VT or VF in the spironolactone group (*p*=0.08).

**Figure 1:**
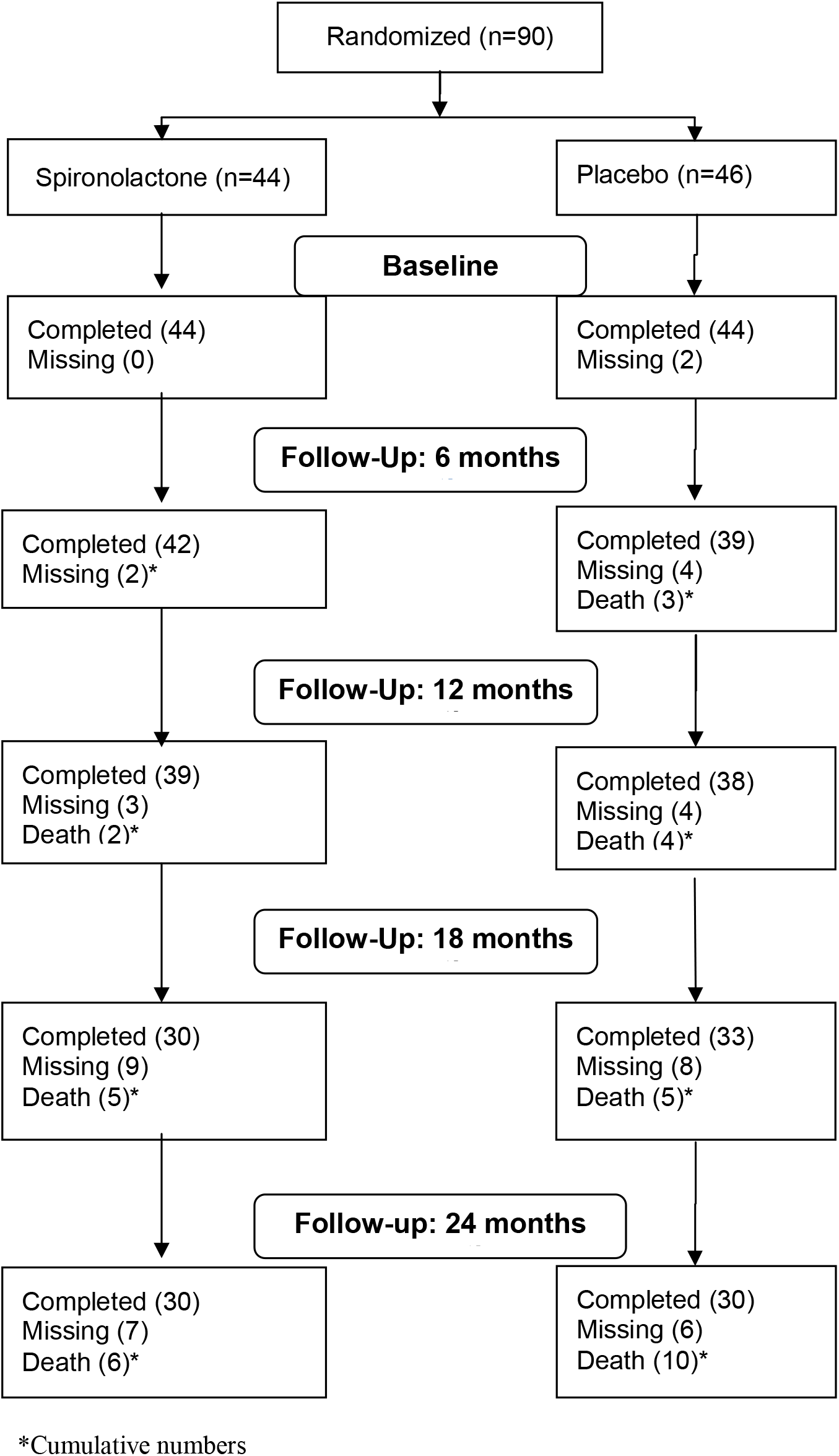
CONSORT diagram for HRQOL outcomes

### HRQOL over 24 months by intent to treat groups

Baseline HRQOL was not statistically different by treatment groups. Additionally, there were no statistically significant differences noted over time in HRQOL between the spironolactone or placebo group with any of the three instruments. HRQOL remained relatively stable over time. This sample reported similar levels of HRQOL to other ICD samples.^13^

### Patient Concerns Assessment (PCA)

Between the two treatment groups, the PCA responses did not significantly change over time in any of the subscales or total scales (Table 3). In general, those in the placebo group initially had a lower number of total concerns than the spironolactone group, and the number of symptoms reported within the placebo group was lower than the spironolactone group over time (F=3.99, p= 0.05). In contrast, patients in the spironolactone group showed more concerns in PCA scales than the placebo group that remained relatively constant over time. The number of symptoms and concerns in this sample is higher than levels reported by others.^14,15^

**Table 3.** Patient Concerns Assessment (PCA) over 24 months by randomization group

| Variable | Group |  |  |  |  |  | Group |  | Time |  | Group x Time |  |
| --- | --- | --- | --- | --- | --- | --- | --- | --- | --- | --- | --- | --- |
|  |  | Baseline* | 6 months | 12 months | 18 months | 24 months | *F* | *p* | *F* | *p* | *F* | *p* |
| Total | Placebo | 18.98 ± 9.95 | 17.82 ± 8.19 | 18.42 ± 10.06 | 19.79 ± 9.56 | 17.93 ± 10.83 | 3.82 | **0.05** | 0.65 | 0.42 | 0.10 | 0.75 |
|  | SPL<sup>†</sup> | 20.0 ± 11.73 | 21.67 ± 12.49 | 22.15 ± 12.28 | 23.15 ± 12.28 | 22.80 ± 12.42 |  |  |  |  |  |  |
| Symptoms | Placebo | 10.91 ± 5.66 | 10.00 ± 4.55 | 10.92 ± 5.69 | 11.76 ± 5.77 | 10.80 ± 6.19 | 3.99 | **0.05** | 2.05 | 0.16 | 0.11 | 0.74 |
|  | SPL<sup>†</sup> | 11.52 ± 6.61 | 12.64 ± 6.95 | 13.10 ± 7.02 | 13.93 ± 6.56 | 13.33 ± 7.42 |  |  |  |  |  |  |
| Fear of Dying | Placebo | 2.61 ± 2.22 | 2.15 ± 1.98 | 1.82 ± 2.05 | 1.88 ± 2.15 | 1.63 ± 2.11 | 2.77 | 0.10 | 0.50 | 0.49 | 0.84 | 0.36 |
|  | SPL<sup>†</sup> | 2.39 ± 2.25 | 2.71 ± 2.43 | 2.54 ± 2.35 | 2.87 ± 2.32 | 2.93 ± 2.41 |  |  |  |  |  |  |
| Distress | Placebo | 4.02 ± 2.72 | 3.77 ± 2.57 | 3.79 ± 2.73 | 4.21 ± 2.55 | 3.73 ± 3.04 | 1.70 | 0.20 | 0.08 | 0.78 | 0.06 | 0.80 |
|  | SPL<sup>†</sup> | 3.96 ± 2.88 | 4.07 ± 2.81 | 4.21 ± 2.81 | 4.40 ± 2.895 | 4.17 ± 2.90 |  |  |  |  |  |  |

<sup>†</sup>Spironolactone.

### Short Form 36 Veterans version (SF-36V)

Between the two treatment groups, SF-36V subscales and component summary scores were not statistically different across time (Table 4). Patients in the spironolactone group reported lower HRQOL compared to those in the placebo group on subscales and summary scores. There were four scales in which the placebo scale scores were higher than that of the spironolactone group. These included: role physical (*p*=0.01), social functioning (p=0.01), role emotional (p=0.05), and mental health (p=0.03). The physical component scores (PCS) did not change over time (p=0.48) nor differ by group (*p*=0.17), but scores were lower than the population mean throughout 24 months of follow-up.^16^ Results were similar for the mental component summary scores. Overall, taking spironolactone did not improve or change HRQOL.

**Table 4.**
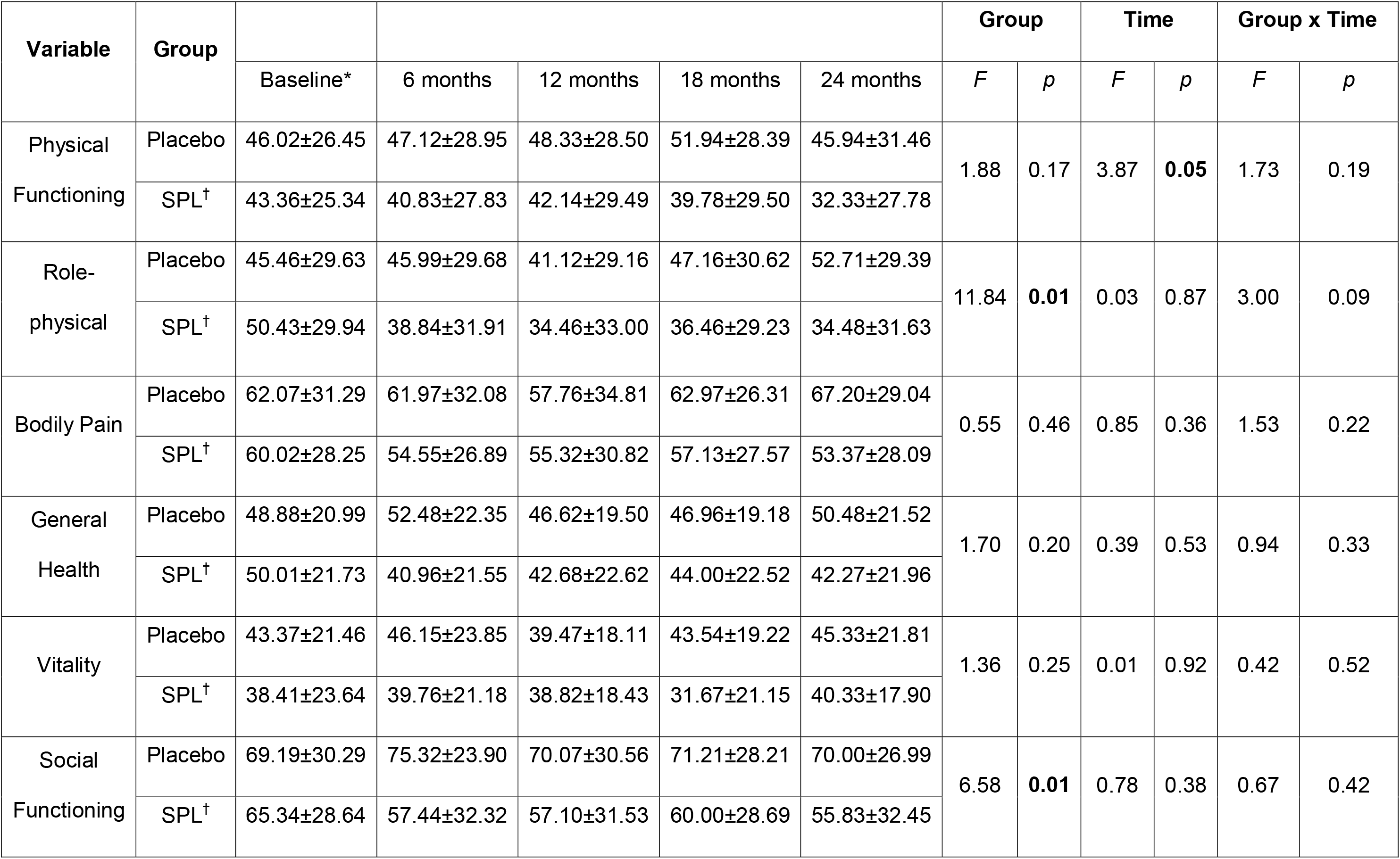
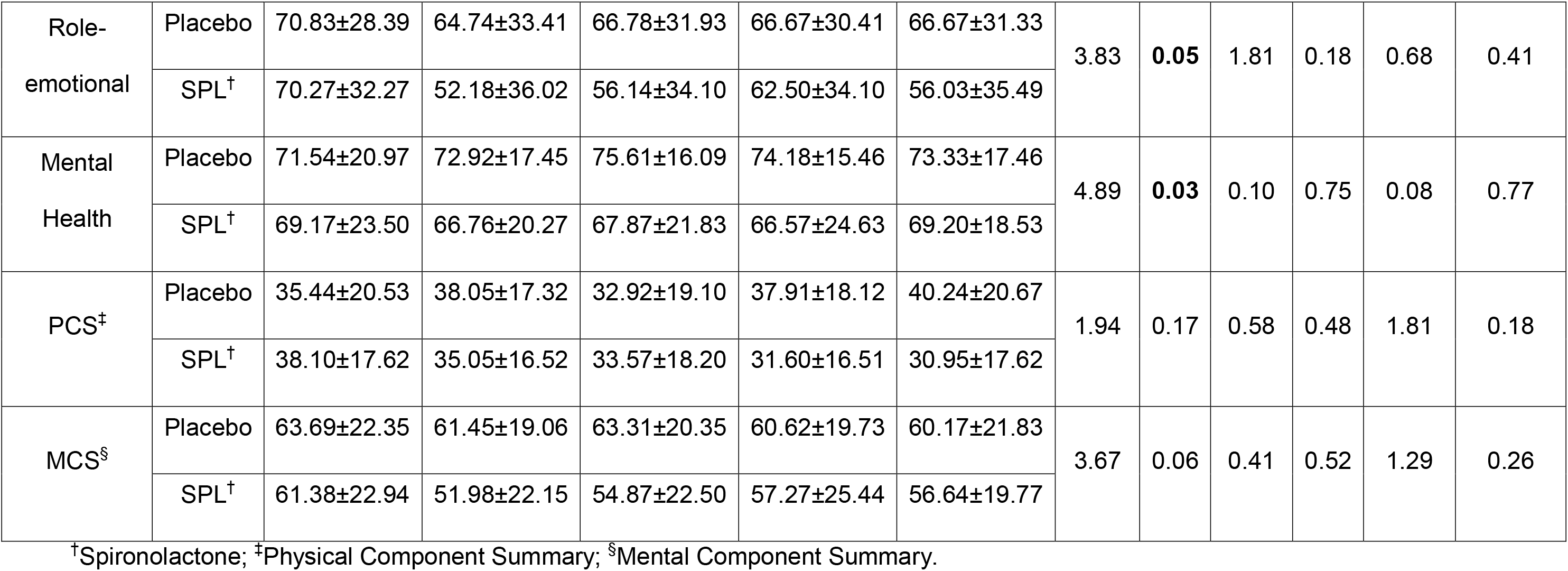
SF-36V over 24 months by randomization group

### Kansas City Cardiomyopathy Questionnaire (KCCQ)

Between the two treatment groups, the KCCQ responses did not significantly change over time in any of the subscales or total scales (Table 5). For both subscales and summary scores of the KCCQ, there were noted significant differences by group such that the placebo group reported higher HRQOL than the spironolactone group over time. This included differences in physical limitation (*p*=0.01), symptom burden (0.02), total symptoms (p=0.04), quality of life (p=0.01), social limitation (p=0.01) and the clinical and overall summary scores. The biggest drop of the scores in the spironolactone group were at the 6-month follow-up and varied with less magnitude during the remainder of the 24-months of follow-up. Conversely, QOL scores in the placebo group were stable over time.

**Table 5.** Kansas City Cardiomyopathy Questionnaire over 24 months by randomization group

| Variable | Group | Occasions |  |  |  |  | Group |  | Time |  | Group x Time |  |
| --- | --- | --- | --- | --- | --- | --- | --- | --- | --- | --- | --- | --- |
|  |  | Baseline* | 6 months | 12 months | 18 months | 24 months | *F* | *p* | *F* | *p* | *F* | *p* |
| Physical limitation | Placebo | 64.43±24.04 | 68.78±22.12 | 64.32±26.86 | 65.73±23.19 | 63.89±27.52 | 6.91 | **0.01** | 6.04 | **0.02** | 0.03 | 0.86 |
|  | SPL<sup>†</sup> | 62.05±27.28 | 55.14±27.34 | 57.28±29.69 | 58.48±26.05 | 47.64±32.58 |  |  |  |  |  |  |
| Symptom stability | Placebo | 50.57±13.73 | 50.00±19.87 | 48.65±16.61 | 50.00±17.96 | 45.83±13.27 | 0.83 | 0.36 | 0.15 | 0.70 | 0.84 | 0.36 |
|  | SPL<sup>†</sup> | 53.98±18.63 | 44.64±14.12 | 49.34±17.90 | 47.50±17.80 | 46.67±20.48 |  |  |  |  |  |  |
| Symptom frequency | Placebo | 84.28±21.39 | 85.20±23.99 | 80.01±25.77 | 80.01±24.30 | 83.54±27.55 | 2.23 | 0.14 | 3.33 | 0.07 | 0.01 | 0.96 |
|  | SPL<sup>†</sup> | 74.34±28.87 | 67.36±31.07 | 69.41±29.37 | 66.46±30.75 | 65.76±29.47 |  |  |  |  |  |  |
| Symptom burden | Placebo | 74.43±20.44 | 75.22±20.09 | 70.27±22.70 | 72.66±21.92 | 75.83±21.26 | 5.59 | **0.02** | 1.21 | 0.275 | 1.08 | 0.30 |
|  | SPL<sup>†</sup> | 67.42±24.89 | 61.31±25.76 | 63.60±24.62 | 58.61±27.20 | 57.78±27.59 |  |  |  |  |  |  |
| Total symptoms | Placebo | 79.36±19.99 | 80.21±21.53 | 75.14±23.78 | 76.33±22.40 | 79.69±23.78 | 4.16 | **0.04** | 2.28 | 0.13 | 0.47 | 0.50 |
|  | SPL<sup>†</sup> | 70.88±26.06 | 64.34±27.66 | 66.50±25.88 | 62.53±28.23 | 61.77±27.63 |  |  |  |  |  |  |
| Quality of Life | Placebo | 65.72±28.20 | 70.62±26.19 | 66.89±26.17 | 69.40±26.60 | 69.44±23.81 | 6.2 | **0.01** | 0.04 | 0.84 | 0.03 | 0.85 |
|  | SPL<sup>†</sup> | 60.80±26.08 | 53.77±29.63 | 59.21±28.98 | 53.89±26.60 | 53.33±32.13 |  |  |  |  |  |  |
| Social<br>Limitation | Placebo | 63.67±28.46 | 67.57±30.47 | 66.12±27.84 | 68.55±26.14 | 65.52±28.77 | 6.35 | **0.01** | 0.01 | 0.99 | 0.34 | 0.56 |
|  | SPL<sup>†</sup> | 56.30±26.87 | 46.83±30.76 | 50.11±33.35 | 48.54±31.02 | 45.19±34.31 |  |  |  |  |  |  |
| Clinical<br>summary | Placebo | 72.14±19.21 | 74.45±19.63 | 69.46±23.12 | 71.42±21.63 | 71.79±24.48 | 5.26 | **0.02** | 5.42 | **0.02** | 0.01 | 0.95 |
|  | SPL<sup>†</sup> | 66.97±25.67 | 59.74±26.67 | 61.03±27.87 | 60.54±25.37 | 55.55±28.58 |  |  |  |  |  |  |
| Overall<br>summary | Placebo | 68.74±21.04 | 71.75±21.88 | 68.67±22.55 | 69.82±22.27 | 69.90±23.45 | 7.77 | **0.01** | 1.81 | 0.18 | 0.07 | 0.79 |
|  | SPL<sup>†</sup> | 62.91±23.35 | 55.02±27.05 | 57.35±28.59 | 55.56±25.46 | 51.96±28.43 |  |  |  |  |  |  |

<sup>†</sup>Spironolactone.

### HRQOL over 24 months by shock versus no-shock groups

When comparing HRQOL between those who experienced at least one ICD shock between the follow-up periods compared to those who had no ICD shocks (Table 6), HRQOL was significantly lower at 6-months on the PCA total score, SF-36V-PCS and MCS scores, and the KCCQ clinical and overall summary scores. Similar group differences in HRQOL between shock and no shock groups were found at 24-months on the PCA (p=0.01) and the KCCQ clinical (p=0.01) and overall summary scores (p=0.005). Receipt of an ICD shock results in significantly lower HRQOL and higher number of reported symptoms.

**Table 6.** Comparison of ICD shock vs. no shock groups: PCA, SF-36V, KCCQ

| Variable | Group | Occasion |  |  |  |  |
| --- | --- | --- | --- | --- | --- | --- |
|  |  | Baseline | 6 months | 12 months | 18 months | 24 months |
| PCA | Shock | 21.59±10.66 | 25.46±11.38 | 21.25±11.96 | 22.43±10.03 | 23.38±12.29 |
|  | No Shock | 18.71±10.87 | 17.61±9.76 | 19.63±10.63 | 20.74±10.45 | 16.8±10.08 |
|  | *F*, P-value* | 1.32, 0.25 | 14.51, **<0.001** | 0.02, 0.88 | 0.82, 0.37 | 9.61, **0.003** |
| SF-36V (PCS) | Shock | 31.91±16.15 | 28.07±10.76 | 28.77±17.25 | 28.77±17.25 | 35.23±18.87 |
|  | No Shock | 38.89±20.16 | 39.61±18.03 | 36.70±19.44 | 36.78±20.50 | 36.76±21.59 |
|  | *F*, P-value* | 2.42, 0.12 | 4.13, **0.05** | 1.83, 0.18 | 1.60, 0.21 | 0.25, 0.62 |
| SF-36V (MCS) | Shock | 63.42±23.79 | 48.59±15.94 | 56.90±20.79 | 57.86±21.95 | 54.80±21.24 |
|  | No Shock | 61.62±22.03 | 60.04±21.98 | 61.98±21.55 | 59.61±23.71 | 63.57±18.01 |
|  | *F*, P-value* | 0.11, 0.75 | 8.66, **0.004** | 1.12, 0.29 | 0.11, 0.74 | 3.02, 0.09 |
| KCCQ clinical summary | Shock | 63.59±20.33 | 51.97±20.11 | 55.20±25.84 | 62.81±22.48 | 56.20 ±26.16 |
|  | No Shock | 68.99±22.49 | 69.40±23.39 | 69.01±23.26 | 65.35±24.24 | 69.34±26.34 |
|  | *F*, P-value* | 1.13, 0.29 | 11.95, **<0.001** | 5.53, **0.02** | 0.20, 0.66 | 6.39, **0.01** |
| KCCQ overall summary | Shock | 60.99±21.46 | 46.92±21.61 | 53.95±27.17 | 60.86±24.83 | 54.51±27.43 |
|  | No Shock | 66.22±22.09 | 67.53±24.70 | 67.68±23.66 | 62.71±24.65 | 67.00±25.32 |
|  | *F*, P-value* | 1.06, 0.31 | 15.35, **<0.001** | 7.26, **0.009** | 0.91, 0.34 | 8.68, **0.005** |

ANOVA controlling for baseline value of the variable with shock vs. no shock group experiencing at least 1 shock before the measurement time; \*p-values are difference between groups (shock and no shocks) by occasion

## DISCUSSION

This study demonstrated that taking spironolactone compared to placebo to delay the recurrence of VT/VF and ICD shocks had no significant improvement or decrement in HRQOL or symptoms over 24 months. The biggest benefit of spironolactone in the treatment of HF on HRQOL in the RALES trial,^3^ was in improved physical wellbeing. Thus, it is not surprising that since RALES eligible patient were excluded from this study, we did not demonstrate a QOL benefit with spironolactone. However, those who received ICD shocks reported significantly lower HRQOL in the measurement period directly after receiving an ICD shock. A temporal association between receipt of an ICD shock and subsequent reductions in HRQOL were evident in both treatment groups.

HRQOL in the Amiodarone vs. Implantable Defibrillator (AVID)^8^ trial demonstrated no significant differences between those receiving an ICD or those receiving amiodarone at 3, 6, and 12 months using the SF-36 and Patient Concerns Assessment (PCA). Physical functioning increased with time in both groups, no trends were noted in mental health scores with time. Patient Concerns in both groups declined with time. Adverse symptoms were related to lower HRQOL scores in both groups. Those receiving ICD shocks reported lower HRQOL and greater numbers of concerns than those not receiving ICD shocks,^8^ similar to our findings.

The SCD-HeFT trial^17^ compared ICD therapy or amiodarone with optimal medical therapy alone in 2,521 patients with moderately symptomatic stable HF, noting that amiodarone had no significant effects on QOL outcomes during 30 months of follow-up. However, ICD shocks within the month preceding a scheduled assessment were associated with decreased HRQOL in multiple domains. Also observed was a temporal association between shocks and HRQOL when comparing shock and no-shock group at 3, 12 and 30 months with baseline scores.

In the Canadian Implantable Defibrillator Study (CIDS),^18^ HRQOL measured at 2, 6, and 12 months was significantly better in those receiving an ICD as compared to those receiving amiodarone using the Mental Health Inventory (MHI) and the Nottingham Health Profile. The amiodarone group demonstrated no improvements or declines in emotional and physical health scores with time. However, HRQOL did not improve in the group who received 5 or more ICD shocks over one year.

For patients who experience ICD shocks, data suggest unchanged or lower HRQOL compared to patients without ICD therapy. A high number of shocks and frequency of shocks is correlated with reduced HRQOL. Investigations suggest this occurs because repeated shock experiences negatively impact perception of health. These results may be confounded with severity of illness, such that those who receive more ICD shocks are in general in poor health.^19,20^ From these early findings, support efforts at reducing ICD shocks through innovative programming,^21^ adjuvant medical therapy,^22^ and ablation^23^ have been reported.

Health care providers are in a key position to assist individuals with making positive adjustments in living successfully with the ICD implantation, including the prevention of as many ICD shocks as possible and the provision for more psychological support to those receiving ICD shocks. Improving HF symptoms and functional class by prudent medical management will perhaps confer improvements to overall HRQOL in those with an ICD. HRQOL is low and symptoms and concerns are high in this population compared to other CV disease populations.^24^ Interventions to address these important patient reported outcomes are warranted.

There are a number of strengths of this study. First, several HRQOL instruments were used to capture the concept of HRQOL, so that differences in measures could be compared over time. Second, this study reinforces the temporal association between HRQOL and ICD shocks, in which those with ICD shocks reported lower HRQOL over time. Third, a longer follow-up time period of 24-months was used, which is longer than most trials in the ICD population. Finally, no patients were lost to follow-up, 14% died in the spironolactone group and 22% in the placebo group over 24 months.

The limitations of this study include the low number of participants that were enrolled overall, and the study was not initially powered to find statistically significant differences in HRQOL. Instead, we aimed to describe the effects of spironolactone on these outcomes. HRQOL was lower in the spironolactone group overall, although there were no statistically significant baseline differences between groups. Finally, the ICD shock vs no shock groups were created after randomization and thus these results should be interpreted conservatively.

## CONCLUSION

The use of spironolactone over a 24-month period did not significantly affect HRQOL or symptoms in patients with an ICD. Patients receiving one or more ICD shocks reported a significant reduction in HRQOL. Longer term HRQOL after receipt of an ICD for secondary prevention of sudden cardiac arrest is stable over time, but lower than other samples of patients with heart disease. Further development of interventions to reduce ICD shocks and address HRQOL in those living with an ICD long term are warranted.

## Data Availability

Data is available upon request.

